# Optimization of Ventilation Therapy Prioritization Strategies among Patients with COVID-19: Lessons Learned from Real-World Data of nearly 600,000 Hospitalized Patients

**DOI:** 10.1101/2022.08.04.22278438

**Authors:** Mohsen Abbasi-Kangevari, Ali Ghanbari, Mohammad-Reza Malekpour, Seyyed-Hadi Ghamari, Sina Azadnajafabad, Sahar Saeedi Moghaddam, Mohammad Keykhaei, Rosa Haghshenas, Ali Golestani, Mohammad-Mahdi Rashidi, Nazila Rezaei, Erfan Ghasemi, Negar Rezaei, Hamid Reza Jamshidi, Bagher Larijani

## Abstract

**Objective:** To investigate the benefit of ventilation therapy among various patient groups with COVID-19 admitted to hospitals, based on the real-world data of hospitalized adult patients.

**Methods:** Data used in the longitudinal study included 599,340 records of hospitalized patients. All participants were categorized based on demographics and their date of hospitalization. Two models were used in this study: firstly, participants were assessed by their probability of receiving ventilation therapy during hospitalization using mixed-effects logistic regression. Secondly, the clinical benefit of receiving ventilation therapy among various patient groups was quantified while considering the probability of receiving ventilation therapy during hospital admission, as estimated in the first model.

**Findings:** Among participants, 60,113 (10.0%) received ventilation therapy, 85,158 (14.2%) passed away due to COVID-19, and 514,182 (85.8%) recovered. Among all groups with sufficient data for analysis, patients aged 40-64 years who had chronic respiratory diseases (CRD) and malignancy benefitted the most from ventilation therapy; followed by patients aged 65+ years who had malignancy, cardiovascular diseases, and diabetes; and patients aged 18-39 years who had malignancy. Patients aged 65+ who had CRD and cardiovascular disease gained the least benefit from ventilation therapy.

**Conclusion:** This study promotes a new aspect of treating patients for ventilators: it could be suggested that rather than focusing on the scarcity of ventilators, guidelines focus on decision-making algorithms to also take the usefulness of the intervention into account, whose beneficial effect is dependent on the selection of the right time in the right patient.

**Funding:** This work was supported by the World Health Organization (WHO) Eastern Mediterranean Regional Office (EMRO) (Grant No. 202693061). The funders had no role in study design, data collection and analysis, decision to publish, or preparation of the manuscript.

**Research in context:** *What was already known:* Research has been ongoing to investigate the main principles for allocating scarce medical resources during pandemics. Medical experts working at the COVID-19 care units interact with patients of different socioeconomic, clinical, paraclinical, and overall health statuses. While physicians should not be faced with situations where they would be obliged to decide which patient to treat due to the risk of human error as well as the double-burden of life-long emotional toll, the pandemic has increased the likelihood of such dilemmas, especially in settings with limited resources. Serious discussions on the ethical considerations of ventilator allocation were also raised during the pandemic. Utility (maximizing benefits) and equity (distributive justice) were two concerns raised in decision making in such dilemma which has also been considered to be “the toughest triage”.

*What new knowledge the manuscript contributes:* This longitudinal study provides new insights on optimizing the strategies for ventilation therapy prioritization among patients with COVID-19, based on the real-world data of nearly 600,000 hospitalized patients with COVID-19. So far, there has been focus on how to prioritize patients with COVID-19 for ventilation therapy. Nevertheless, there has not been much evidence on how much patients of different age groups with various underlying conditions actually benefitted from ventilation therapy based on real-world data. The results of this study could have a significant message: should the prioritization guidelines for ventilators allocation take no notice of the real-world data, patients might be deprived of ventilation therapy, who could benefit the most from it. This would pave the way to capture clearer picture in the possible future pandemics.

## Background

The coronavirus disease-2019 (COVID-19) pandemic has far officially claimed more than 5.22 million lives worldwide (1). Acute respiratory distress syndrome (ARDS), a common complication of COVID-19 among critically ill patients, requires medical management involving ventilation therapy. Of all patients diagnosed with COVID-19, 17% to 35% would be hospitalized at intensive care units (ICUs) (2,3), and 9% to 19% would require invasive mechanical ventilation (2,4). The availability of ICU beds varies widely between countries, even among the wealthiest countries (5). While COVID-19 continues to place extraordinary demands on healthcare systems, resulting in severe shortages of essential resources and services (6), the scarcity of ventilators could be the most challenging, as there is typically limited time if mechanical ventilation is vital (7). The estimated number of available invasive mechanical ventilators in various countries would not be adequate to serve all clinically eligible patients during the pandemic (8).

Research has been ongoing to investigate the main principles for allocating scarce medical resources during pandemics (9–11). Medical experts working at the COVID-19 care units interact with patients of different socioeconomic, clinical, paraclinical, and overall health statuses. While physicians should not be faced with situations where they would be obliged to decide which patient to treat due to the risk of human error as well as the double-burden of life-long emotional toll, the pandemic has increased the likelihood of such dilemmas, especially in settings with limited resources (12). Thus, prioritization recommendations and guidelines are under development in the hope of helping physicians, especially those less experienced, with the real-time decision-making process based on the resources and contexts (6,13). Serious discussions on the ethical considerations of ventilator allocation were also raised during the pandemic. Utility (maximizing benefits) and equity (distributive justice) were two concerns raised in decision making (14,15) in such dilemma which has also been considered to be “the toughest triage” (7). From a utilitarian perspective, saving the most lives or saving the most life-years by allocation of ventilation to those with higher survival could guide rationing (7,14,15).

Nevertheless, there is not much information about ventilation therapy for patients with COVID-19. Drawing from previous World Health Organization (WHO) guidelines, there are recommendations to indicate which patients with hypoxemic respiratory failure should be considered for non-invasive ventilation and prioritize in settings with limited resources (16). It remains challenging yet imperative to prioritize therapy to patients who will benefit the most from it considering availability and risk, considering the increased risk of infection transmission when the patient undergoes endotracheal intubation and non-invasive ventilation (17). Determining which patients with COVID-19 would benefit the most from ventilation therapy could help optimize the current ventilator allocation guidelines. Thus, the objective of this study was to investigate the benefit of ventilation therapy among various patient groups with COVID-19 admitted to hospitals, based on the real-world data of hospitalized adult patients.

## Material and methods

### Ethics

This work was supported by the WHO EMRO Office (EMRO) (Grant No. 202693061). The ethics committee of Endocrinology and Metabolism Research Center, Endocrinology and Metabolism Clinical Sciences Institute, Tehran University of Medical Sciences, Tehran, Iran, approved this study under the reference number IR.TUMS.EMRI.REC.1400.034. The confidentiality of the data and the results are preserved.

### Overview

Data used included 599,340 records of hospitalized patients with COVID-19 in Iran who were admitted from February 2020 to June 2021. Patients were categorized based on sex, age, city of residence, the hospitals’ affiliated university, date of hospitalization, and comorbidities. First, the probability of patients’ ventilation therapy during hospitalization was calculated. Then, patients’ survival was assessed and the clinical benefit of ventilation therapy among various patient groups was quantified while considering the probability of receiving ventilation therapy during hospital admission, as estimated in the first model.

### Data source and variables

Data of this longitudinal study were retrieved from the Iranian COVID-19 Registry provided by the Ministry of Health and Medical Education, which was gathered from hospitals and included patients with COVID-19 in Iran from the early days of the pandemic. Data used in the current study included 599,340 records of hospitalized patients who were admitted from February 2020 to June 2021. The study variables included the patients’ age; sex; underlying conditions, including diabetes mellitus (DM), cardiovascular diseases (CVD), chronic respiratory disease (CRD), malignancy; receiving ventilation therapy; and COVID-19 outcomes, including recovery or death.

### Case definitions

DM, CVD, CRD, and malignancy were obtained from patients’ self-reported medical history. The diagnosis of COVID-19 was made by physicians based on a positive Real-Time Reverse

Transcription Polymerase Chain Reaction (RT-PCR) result for SARS-CoV-2, or clinical suspicion defined as (1) at least two of the following symptoms lasting for at least 48 hours: fever (axillary temperature ≥37.5°C), chills, sore throat, stuffy nose, myalgia, fatigue, headache, nausea or vomiting, or diarrhea or (2) at least one respiratory sign or symptom (including cough, shortness of breath), new olfactory or taste disorder, or radiographic evidence of COVID-19–like pneumonia.

## Data analysis

### Variables

All participants were categorized based on sex, age, city of residence, the hospitals’ affiliated university, and their date of hospitalization. Age groups were defined as 18-39, 40-64, and more than 65-year-old participants. The affiliated university were assessed due to the possibility of using disparate approaches and guidelines regarding ventilator allocation policies. The date of hospitalization was also included due to the paramount importance of considering the scarcity of vital equipment at the peak of the COVID-19 epidemic surge. The intervals included in the analysis were as follows: February-March 2020, April-May 2020, June-July 2020, August-September 2020, November-December 2020, January-February 2021, March-April 2021, and May-June 2021. In addition to demographic annotations, patients’ data were further assessed for comorbidities and underlying/clinical conditions, which included CRD, CVD, DM, and malignancies.

### Statistical methods

Two models were used in this study: in the first model, participants were assessed by their probability of receiving ventilation therapy during hospitalization based on demographic and clinical factors using mixed-effects logistic regression. In the second model, the clinical benefit of receiving ventilation therapy among various patient groups was quantified while considering the probability of receiving ventilation therapy during hospital admission, as estimated in the first model.

### Estimating the probability of ventilation therapy

First, we used a mixed-effects logistic regression model (18) to estimate the probability of receiving ventilation therapy among patients. The response variable was binary, with “one” representing receiving ventilation therapy. The effects of time intervals, age groups and affiliated university were considered as random intercept effects. Sex, ICU admission, CRD, malignancy, CVD, and diabetes were random intercept effects that varied among different age groups, as presented in the following:

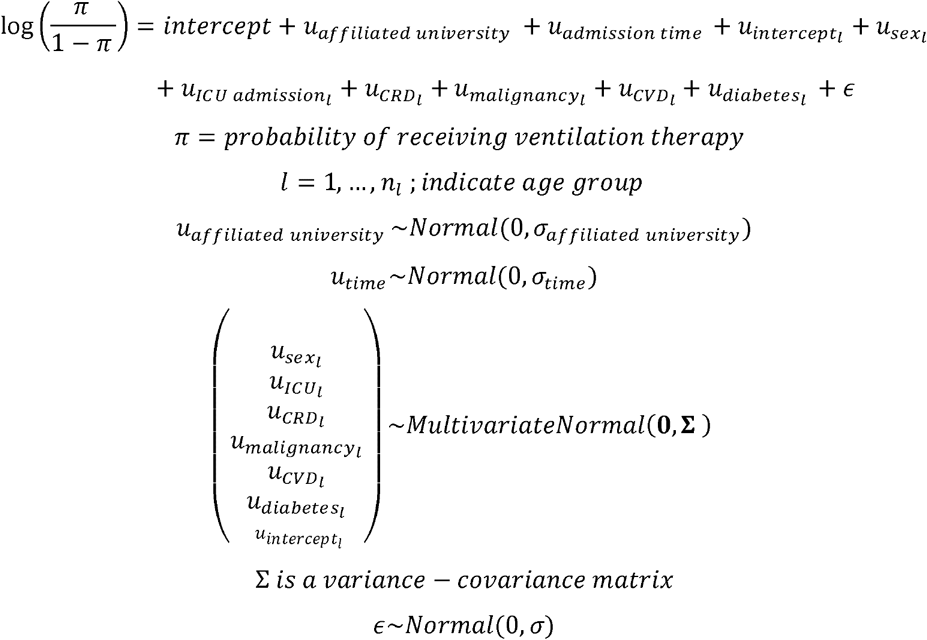

### Estimating the probability of recovery

To investigate the extent of benefit among patients with various underlying conditions, the uneven chance of receiving ventilation therapy due to the time of admission, hospital equipment, or resource allocation guidelines used needed to be addressed. First, we divided the patients into 48 groups based on their age groups and underlying conditions, including CRD, malignancy, CVD, DM. Then, considering the high sample size and to simplify the modeling process, a logistic generalized linear model was fitted separately for each group. The response variable was binary with “one” representing recovery. Also, the admission province, admission time, patient sex, and ICU admission were the independent variables. The last term of the model was the interaction between a binary variable, with “one” representing receiving ventilation therapy, and a continuous variable indicating the probability of receiving ventilation therapy obtained from the first model. This interaction gives away two main effects and one interaction coefficient, as presented in the following:

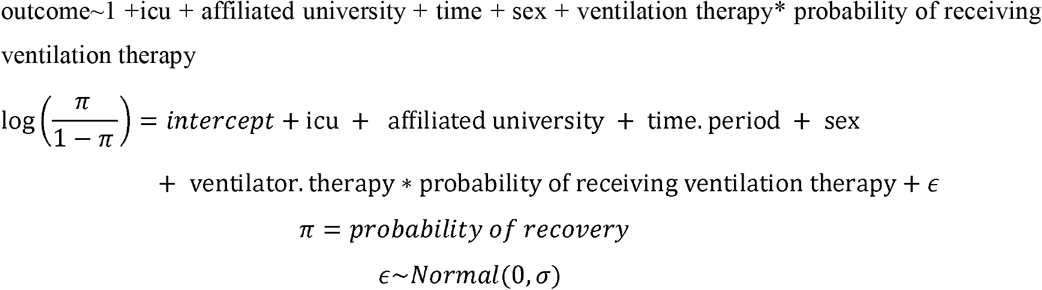

### Main effects

The first main effect indicated the ratio of the odds for recovery among patients who received ventilation therapy to the odds for those who did not, while considering other factors constant. The second main effect indicated the ratio of the odds for recovery for a one-unit increase (“zero” probability represents not receiving ventilation therapy, while “one” represents receiving ventilation therapy) in the probability of receiving ventilation therapy, while considering other factors constant.

### Interaction coefficient

The interaction coefficient indicated the difference in the slope of the logit probability of recovery for a one-unit increase in the probability of receiving ventilation therapy between the patients who received ventilation compared to those who did not while considering other factors constant. We considered the positive and significant coefficient values to represent the benefit of receiving ventilation for patients who receive ventilation compared to those who did not. Also, a higher value of this coefficient indicated more benefit. The interaction coefficient could be used as an indicator to quantify the benefit of ventilation reception and possibly be used as a criterion for comparison among various patient groups.

## Results

Data of 599,340 participants were analyzed, which encompassed 60,113 (10.0%) cases with ventilation therapy, 85,158 (14.2%) cases who died, and 514,182 (85.8%) cases who recovered. The mean (SD) age was 58. 5 (18.3) [range= 18-114, being 58.3 (18.2) among women, and 58.6 (18.4) among men]. Characteristics of participants are presented in Table 1. The COVID-19 outcome based on sex, age-groups and underlying diseases are presented in Figure 1.

**Table 1.** Characteristics of participants

| Variable | N (%) |
| --- | --- |
| ***Demographics*** |  |
| Sex |  |
| Female | 291,267 (48.6) |
| Male | 308,073 (51.4) |
| Age |  |
| 18-39 years | 113,211 (18.9) |
| 40-64 years | 240,298 (40.1) |
| More than 65 years | 245,831 (41.0) |
| ***Underlying conditions*** |  |
| CVD |  |
| Yes | 110,593 (18.5) |
| No | 488,747 (81.5) |
| DM |  |
| Yes | 84,973 (14.2) |
| No | 514,367 (85.8) |
| COPD |  |
| Yes | 26,153 (4.4) |
| No | 573,187 (95.6) |
| Malignancy |  |
| Yes | 10610 (2.0) |
| No | 588730 (98.0) |
| ***Outcomes*** |  |
| Ventilation therapy | 60,113 (10.0) |
| Death | 85,158 (14.2) |
| Recovery | 514,182 (85.8) |

**Figure 1.**
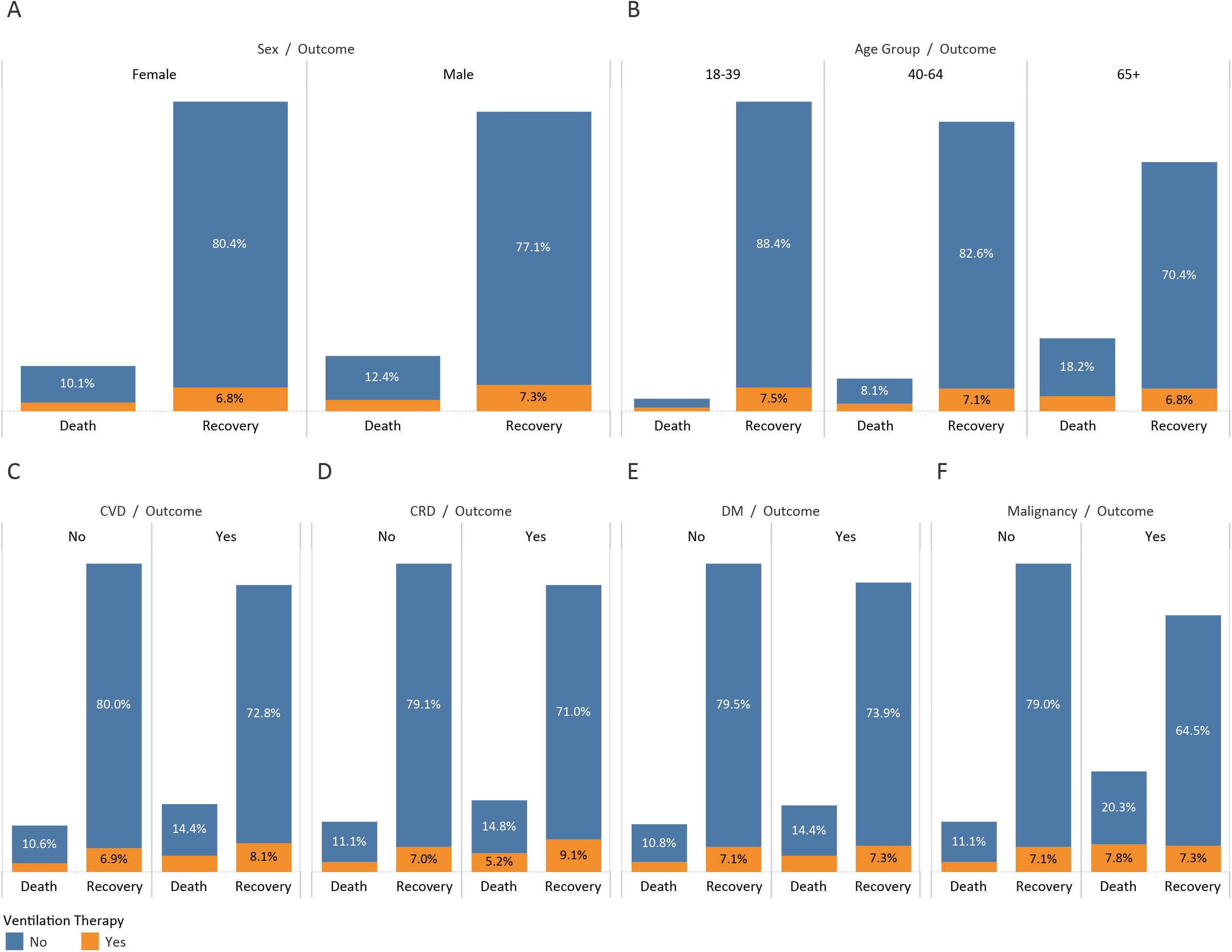
COVID-19 outcome based on (A) sex, (B) cardiovascular disease, (C) chronic respiratory disease, (D) malignancy, (E) diabetes mellitus, and (F) age groups.

Among all combinations, analysis of eight groups was not available due to paucity of data (Table 2). Among all groups with sufficient data for analysis, patients aged 40-64 years who had CRD and malignancy benefitted the most from ventilation therapy; followed by patients aged 65+ years who had malignancy, cardiovascular diseases, and diabetes; and patients aged 18-39 years who had malignancy. Patients aged 65+ who had CRD, and cardiovascular disease gained the least benefit from ventilation therapy. Among patients with diabetes, patients aged 65+ years benefited from ventilation therapy, followed by 40-64 years. Among patients with cardiovascular diseases, patients aged 18-39 years benefited the most from ventilation therapy, followed by patients aged 40-64 years and 65+ years. Among patients with diabetes and cardiovascular diseases, patients aged 40-64 years benefited from ventilation therapy, followed by 65+ years. Among patients with no history of CRD, malignancy, cardiovascular disease, or diabetes, patients aged 18-39 years benefited the most from ventilation therapy, followed by patients aged 40-64 years and 65+ years (Figure 2).

**Table 2.** The estimated benefit of ventilation therapy among patients of different age groups with various underlying conditions

| id | COPD | Malignancy | CVD | DM | Age | Benefit * | p-value | Observation<br>(Total<sup>**</sup>) | Observation<br>(Ventilation<sup>***</sup>) | Observation<br>(Recovery<sup>****</sup>) |
| --- | --- | --- | --- | --- | --- | --- | --- | --- | --- | --- |
| 1 | No | No | No | No | 18-39 | 1.983 | <0.001 | 103991 | 8574 | 100269 |
| 2 | No | No | No | No | 40-64 | 1.728 | <0.001 | 171041 | 14566 | 155469 |
| 3 | No | No | No | No | 65+ | 1.369 | <0.001 | 139453 | 14014 | 108579 |
| 4 | No | No | No | Yes | 18-39 | 1.267 | N/S | 2827 | 288 | 2654 |
| 5 | No | No | No | Yes | 40-64 | 1.455 | <0.001 | 22096 | 2214 | 19465 |
| 6 | No | No | No | Yes | 65+ | 1.72 | <0.001 | 23258 | 2705 | 17752 |
| 7 | No | No | Yes | No | 18-39 | 1.836 | 0.008 | 2895 | 336 | 2667 |
| 8 | No | No | Yes | No | 40-64 | 1.31 | <0.001 | 22712 | 2539 | 20251 |
| 9 | No | No | Yes | No | 65+ | 1.266 | <0.001 | 43934 | 5722 | 34249 |
| 10 | No | No | Yes | Yes | 18-39 | -2.412 | N/S | 401 | 46 | 361 |
| 11 | No | No | Yes | Yes | 40-64 | 1.877 | <0.001 | 10660 | 1276 | 9088 |
| 12 | No | No | Yes | Yes | 65+ | 1.427 | <0.001 | 19903 | 2598 | 15205 |
| 13 | No | Yes | No | No | 18-39 | 3.639 | <0.001 | 1136 | 136 | 858 |
| 14 | No | Yes | No | No | 40-64 | 1.255 | 0.004 | 3788 | 528 | 2816 |
| 15 | No | Yes | No | No | 65+ | 0.691 | N/S | 3085 | 501 | 2098 |
| 16 | No | Yes | No | Yes | 18-39 | N/A | N/A | 16 | 2 | 15 |
| 17 | No | Yes | No | Yes | 40-64 | -0.324 | N/S | 313 | 44 | 242 |
| 18 | No | Yes | No | Yes | 65+ | 2.592 | N/S | 388 | 72 | 258 |
| 19 | No | Yes | Yes | No | 18-39 | N/A | N/A | 21 | 6 | 16 |
| 20 | No | Yes | Yes | No | 40-64 | -0.808 | N/S | 234 | 44 | 181 |
| 21 | No | Yes | Yes | No | 65+ | 1.867 | N/S | 617 | 111 | 440 |
| 22 | No | Yes | Yes | Yes | 18-39 | N/A | N/A | 7 | 1 | 6 |
| 23 | No | Yes | Yes | Yes | 40-64 | 4.885 | N/S | 132 | 16 | 99 |
| 24 | No | Yes | Yes | Yes | 65+ | 3.79 | 0.049 | 279 | 45 | 206 |
| 25 | Yes | No | No | No | 18-39 | 0.499 | N/S | 1685 | 238 | 1582 |
| 26 | Yes | No | No | No | 40-64 | 0.873 | 0.041 | 5958 | 721 | 5204 |
| 27 | Yes | No | No | No | 65+ | 0.466 | N/S | 7095 | 1015 | 5391 |
| 28 | Yes | No | No | Yes | 18-39 | 241.684 | N/S | 44 | 8 | 40 |
| 29 | Yes | No | No | Yes | 40-64 | 3.137 | 0.005 | 834 | 123 | 696 |
| 30 | Yes | No | No | Yes | 65+ | 0.605 | N/S | 1295 | 172 | 977 |
| 31 | Yes | No | Yes | No | 18-39 | 445.595 | N/S | 131 | 14 | 118 |
| 32 | Yes | No | Yes | No | 40-64 | 0.651 | N/S | 1599 | 221 | 1348 |
| 33 | Yes | No | Yes | No | 65+ | 0.76 | 0.032 | 4488 | 730 | 3329 |
| 34 | Yes | No | Yes | Yes | 18-39 | N/A | N/A | 21 | 3 | 18 |
| 35 | Yes | No | Yes | Yes | 40-64 | 0.934 | N/S | 698 | 105 | 572 |
| 36 | Yes | No | Yes | Yes | 65+ | 2.473 | <0.001 | 1711 | 277 | 1274 |
| 37 | Yes | Yes | No | No | 18-39 | -383.91 | N/S | 34 | 6 | 25 |
| 38 | Yes | Yes | No | No | 40-64 | 6.504 | 0.017 | 183 | 31 | 120 |
| 39 | Yes | Yes | No | No | 65+ | -0.061 | 0.982 | 182 | 29 | 120 |
| 40 | Yes | Yes | No | Yes | 40-64 | N/A | N/A | 14 | 1 | 11 |
| 41 | Yes | Yes | No | Yes | 65+ | -338.243 | 1.00 | 31 | 4 | 22 |
| 42 | Yes | Yes | Yes | No | 18-39 | N/A | N/A | 1 | 0 | 1 |
| 43 | Yes | Yes | Yes | No | 40-64 | -6.389 | 1.00 | 24 | 5 | 15 |
| 44 | Yes | Yes | Yes | No | 65+ | -3.169 | 0.494 | 80 | 16 | 49 |
| 45 | Yes | Yes | Yes | Yes | 18-39 | N/A | N/A | 1 | 1 | 1 |
| 46 | Yes | Yes | Yes | Yes | 40-64 | N/A | N/A | 12 | 2 | 9 |
| 47 | Yes | Yes | Yes | Yes | 65+ | 1466.119 | 0.999 | 32 | 7 | 16 |
| *The interaction coefficient of the second model<br>**The total number of patients with COVID-19 who were admitted to the hospital in each group<br>***The total number of patients with COVID-19 who received ventilation therapy in each group<br>****The total number of patients with COVID-19 who recovered after hospital admission in each group |  |  |  |  |  |  |  |  |  |  |

**Figure 2.**
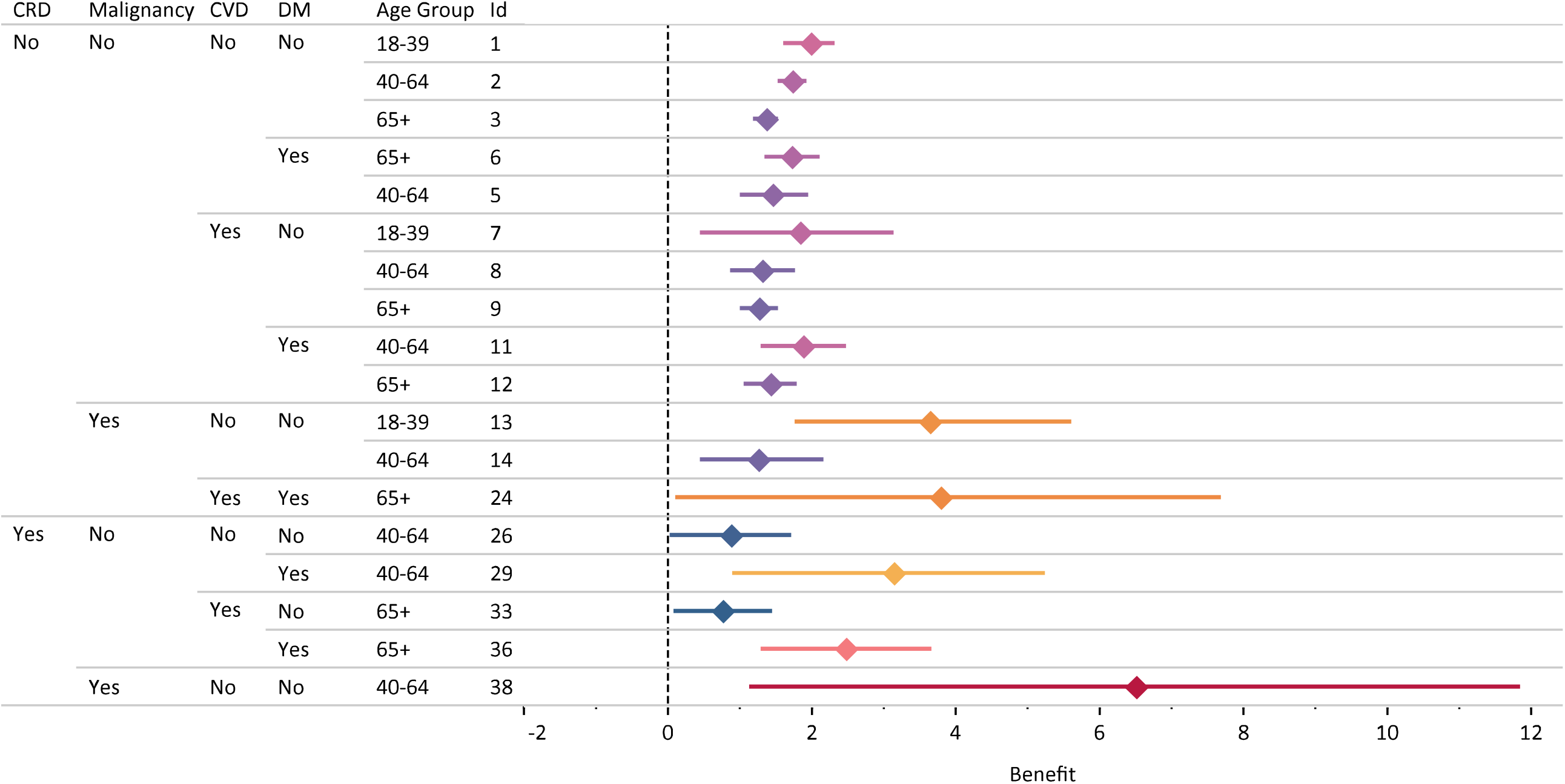
Patient groups who significantly benefited from ventilation therapy: estimated benefit (95% Confidence Interval)

## Discussion

This longitudinal study provides new insights on optimizing the strategies for ventilation therapy prioritization among patients with COVID-19, based on the real-world data of nearly 600,000 hospitalized patients with COVID-19. So far, there has been focus on how to prioritize patients with COVID-19 for ventilation therapy (19). Nevertheless, there has not been much evidence on how much patients of different age groups with various underlying conditions actually benefitted from ventilation therapy based on real-world data. Some studies made endeavours to predict COVID-19 severity (20) or the need for mechanical ventilation (21); however, their approaches have not been investigated in the real-world to determine their outcomes.

In this study, patients aged 40-64 years who had CRD and malignancy benefitted the most from ventilation therapy, followed by patients aged 65+ years who had malignancy, CVD, and DM; and patients aged 18-39 years who had malignancy. Considering that these patient groups are considered to be at moderate or high risk of severe COVID-19 and possibly require ventilation therapy (22), it was propitious that ventilation therapy could increase their chance of recovery.

Patients with COVID-19 who have DM are more likely to require mechanical ventilation (23). Among patients with DM, older age is associated with worse COVID-19 outcomes (24,25). In this study, patients with DM aged 65+ years benefited from ventilation therapy more than those aged 40-64. It is worth mentioning that all patients aged 40+ who only had DM benefitted from ventilation therapy.

Pre-existing CVD is independently associated with COVID-19 adverse outcomes (26). Among patients who only had a CVD in this study, the younger the patients, the more they benefitted from ventilation therapy, unlike what was witnessed for DM. The same age pattern was also seen among patients who had DM and CVD. Some guidelines include age group as an additional consideration (19).

Individuals also usually prioritize younger patients in situations of absolute scarcity of life sustaining resources; however, simply excluding patients from prioritization solely based on their age could be ethically unjustified and biased against older adults (27). Although age-based discrimination includes moral conflicts and socio-cultural issues, ageism has become more apparent since the beginning of the COVID-19 pandemic. The media has played a significant role in this sense, while broadcasting discussions on the age limits for intensive care and ventilation allocation, unintentionally implying that an older person’s life is worth less than a young person’s (28). In this study, among otherwise healthy patients, patients aged 18-39 years benefited the most from ventilation therapy, followed by patients aged 40-64 years, and patients aged 65+ years. While COVID-19 has resulted in severe shortage of ventilators (6) worldwide, countries with limited resources face the most challenges to serve all clinically eligible patients during the pandemic (8). In this sense, factoring the level of benefit each patient would receive from ventilation therapy could help optimizing current guidelines.

In a study, the public opinion on priorities towards the fair allocation of ventilators during the COVID-19 pandemic was investigated, where people assigned a high priority score to patients with underlying diseases (29). This could imply that people assumed that ventilation therapy would generally improve the outcome for patients with underlying conditions. Nevertheless, the real-world data suggested that patients’ age group and underlying diseases could play a significant role in the outcome of ventilation therapy. This calls for knowledge translation by public health authorities and the media to regularly convey the prognostic factors of COVID-19 based on emerging evidence to justify people’s expectations from the healthcare systems

In a Delphi study, a panel of experts were asked to prioritise the allocation of ventilators based on various medical or non-medical factors. While the panel considered patients with active-malignancy to have low priority in receiving ventilation therapy, the real-world data made it crystal clear that patients with malignancy could also benefit from ventilation therapy. Moreover, the panel did not reach a consensus regarding underlying diseases (30). The deviation of real-world data from the experts’ perspectives highlights the potential bias the physicians could have when making a death-life decision, which needs to be taken into account by future guidelines on the fair allocation of ventilators.

Some guidelines assess patients based on their clinical condition at admission, which could include assessment of irreversible shock, and mortality risk using the Sequential Organ Failure Assessment (SOFA) score (31). They also recommend continuous evaluation for withdrawing patients whose clinical condition is not improving despite ventilation therapy (7). Nevertheless, few studies have assessed the application of the current triage criteria to actual patients. In the early days of the pandemic, a retrospective cohort study highlighted how divergent even supposedly similar triage approaches could be, suggesting that different triage approaches identified substantially other patients for initial consideration for withholding or early withdrawal of mechanical ventilation (32). We did not find any studies that investigated the role of ventilation therapy in improving the course of COVID-19 in a setting where patients have been triaged based on SOFA scores.

## Strengths and limitations

This is the first nationwide study to quantify the benefit of ventilation therapy based on the real-world data around 600,000 hospitalized patients of various age groups with COVID-19 who had DM, CVD, malignancy, or CRD. The strength of this study lies in a large sample and data gathering since the early days of the outbreak in Iran. Findings could empower public health authorities to optimize the ventilation therapy prioritization strategies among patients with COVID-19 admitted to hospitals, especially considering that there are currently no national guidelines for allocation of ventilators at the time of resources scarcity in Iran and the decision to prioritize patients for ventilator allocation is performed based on hospital regulations. We realize the limitations of the study. Due to the lack of a national integrated electronic health records system in Iran, many underlying conditions or baseline data of patients, such as their body mass index or behavioral risk factors, were not properly recorded in the COVID-19 registry. Despite the large study population, data points for some patient groups were insufficient for analysis, which need to be addressed in future studies.

## New insights and conclusion

The results of this study could have a significant message: should the prioritization guidelines for ventilators allocation take no notice of the real-world data, patients might be deprived of ventilation therapy, who could benefit the most from it. The comparison of real-world evidence with the general population’s attitudes and medical experts showed an unexpected bias against older age groups and underlying conditions. This study promotes a new aspect of treating patients for ventilators as a scarce medical resource, considering whether ventilation therapy would improve the patient’s clinical outcome. This gains significance considering the divergent outcomes of existing guidelines, especially for patients meeting the lowest priority criteria for mechanical ventilation (32). As a rapidly evolving crisis, numerous therapeutic or preventive approaches are being investigated to lessen the burden of the COVID-19 pandemic (33,34). It could be suggested that rather than focusing on the scarcity of ventilators, guidelines focus on evidence-based decision-making algorithms to also take the usefulness of the intervention into account, similar to some other medications, whose beneficial effect is dependent on the selection of the right time in the right patient (35).

## Data Availability

De-identified, individual participant data will be made available upon requests directed to the corresponding author; after the approval of a proposal, data can be shared through a secure online platform.

## Acknowledgement

We thank all of our colleagues at the Non-Communicable Diseases Research Center (NCDRC), Endocrinology and Metabolism Population Sciences Institute, Tehran University of Medical Sciences, and the supporting roles in Ministry of Health and Medical Education (MOHME) of Iran and WHO Eastern Mediterranean Regional Office (EMRO) who made the conduction of this study possible. The authors would like to appreciate the invaluable contribution of Professor Farshad Farzadfar to this project. They would also like to express their most sincere gratitude to all frontline healthcare workers across the globe during this pandemic.

## Ethical issues

The ethics committee of Endocrinology and Metabolism Research Center, Endocrinology and Metabolism Clinical Sciences Institute, Tehran University of Medical Sciences, Tehran, Iran, approved this study under the reference number IR.TUMS.EMRI.REC.1400.034. The confidentiality of the data and the results are preserved.

